# Just because you’re paranoid doesn’t mean they won’t side with the plaintiff: Examining perceptions of liability about AI in radiology

**DOI:** 10.1101/2024.07.30.24311234

**Authors:** Michael H. Bernstein, Brian Sheppard, Michael A. Bruno, Parker S. Lay, Grayson L. Baird

## Abstract

**Background:** Artificial Intelligence (AI) will have unintended consequences for radiology. When a radiologist misses an abnormality on an image, their liability may differ according to whether or not AI also missed the abnormality.

**Methods:** U.S. adults viewed a vignette describing a radiologist being sued for missing a brain bleed (N=652) or cancer (N=682). Participants were randomized to one of five conditions. In four conditions, they were told an AI system was used. Either AI agreed with the radiologist, also failing to find pathology (AI agree) or did find pathology (AI disagree). In the AI agree+FOR condition, AI agreed with the radiologist and an AI false omission rate (FOR) of 1% was presented. In the AI disagree+FDR condition, AI disagreed and an AI false discovery rate (FDR) of 50% was presented. There was also a no AI control condition. Otherwise, vignettes were identical. Participants indicated whether the radiologist met their duty of care as a proxy for whether they would side with defense (radiologist) or plaintiff in trial.

**Results:** Participants were more likely to side with the plaintiff in the AI disagree vs. AI agree condition (brain bleed: 72.9% vs. 50.0%, p=0.0054; cancer: 78.7% vs. 63.5%, p=0.00365) and in the AI disagree vs. no AI condition (brain bleed: 72.9% vs. 56.3%, p=0.0054; cancer: 78.7% vs. 65.2%, p=0.00895). Participants were less likely to side with the plaintiff when FDR or FOR were provided: AI disagree vs AI disagree+FDR (brain bleed: 72.9% vs. 48.8%, p=0.00005; cancer: 78.7% vs. 73.1%, p=0.1507), and AI agree vs. AI agree+FOR (brain bleed: 50.0% vs. 34.0%, p=0.0044; cancer: 63.5% vs. 56.4%, p=0.1085).

**Discussion:** Radiologists who failed to find an abnormality are viewed as more culpable when they used an AI system that detected the abnormality. Presenting participants with AI accuracy data decreased perceived liability. These findings have relevance for courtroom proceedings.

## Introduction

Advancements in artificial intelligence (AI) for radiology are promising. Van Leeuwen and colleagues^1^ note that AI has the potential to improve patient care through six different clinical objectives, including earlier disease detection and improved diagnostic accuracy. Despite AI’s potential, it is not a panacea, and there will be some unintended consequences.^2^ For instance, one study^3^ showed that radiologists could be misled by inaccurate AI feedback; radiologists were less accurate interpreting the same cases with incorrect AI compared to no AI. Moreover, the study found that radiologists were more likely to be misled when they believed AI results were (versus were not) going into the patient’s file.

Another study (under review) showed that even accurate AI algorithms can have drawbacks. A large vessel occlusion (LVO) detection algorithm was employed with a sensitivity of 100% and specificity of 92%— a very accurate algorithm. However, the false discovery rate (FDR) was 67%—that is, 67% of the cases flagged by AI were false positives. The high FDR was due to the fact LVO had a very low base rate of about 4.0%, resulting in a positive predictive value (PPV) of 33%. Despite the excellent accuracy, the algorithm was nonetheless halted because radiologists perceived that there were too many false positives. Upon surveying the radiologists, most believed the algorithm increased their risk of medical legal liability.

These two studies indicate that when AI results are documented in a patient’s file, radiologists may be apprehensive to disagree with an algorithm when the algorithm flags a case as abnormal. This response is consistent with *adaptive bias*, a concept drawn from cognitive and decision science that describes a desire to minimize the cost of errors rather than the number of errors.^4^ In this context, when AI determines a case is abnormal, the cost of incorrectly disagreeing with AI (false negative) far outweighs the cost of incorrectly agreeing with it (false positive). Banja and colleagues^5^ suggests that a radiologist is more vulnerable to malpractice allegations when they disagree with AI in a false negative case and the patient suffers a poor outcome. By comparison, the consequence of false positives *for a radiologist* is trivial.

The purpose of the present study is to examine this theory empirically. It employs two (brain bleed or lung cancer, respectively) malpractice vignettes where a patient suffers a poor outcome (permanent disability or death, respectively) across five experimental conditions: 1.) no AI, 2.) AI finds pathology, 3.) AI fails to find pathology, 4.) AI finds pathology and the AI’s FDR is reported, 4.) AI fails to find pathology and the AI’s false omission rate (FOR) is reported.

We hypothesized that mock jurors would be more likely to side with a plaintiff (i.e. against the defense, the radiologist) in a malpractice lawsuit in the AI disagree condition versus the AI agree. We further hypothesized that people would be more likely to side with the plaintiff in the AI disagree condition versus the AI disagree + FDR condition – that is, we expected that presenting the false discovery rate would reduce the perceived culpability of a radiologist when a radiologist disagrees with AI. Last, we hypothesized that people would be more likely to side with the plaintiff in the AI agree condition versus the AI agree + FOR condition – that is, we expected that presenting the false omission rate would reduce the perceived culpability of a radiologist when a radiologist agrees with AI. Finally, all AI conditions will be compared with a no AI control condition.

## Materials and Methods

### Participants

Participants were recruited using the online recruitment platform Prolific. Participants were eligible if they were English speaking adults in the U.S. aged 18–89-year-olds. We excluded n=166 and n=135 participants for failing at least one manipulation or attention check item in the brain bleed and cancer vignettes, respectively.

Participants in the brain bleed vignette (n=652) were 53.2% female, 68.6% White/Caucasian, and 18-78 years old (median=44) with a median annual household income of $68,000 (range:0-600,000). Participants in the cancer vignette (n=682) were 54.8% female, 72.7% White/Caucasian, and 18-85 years old (Median=45) with a median annual household income of $68,000 (range: 0-875,000). See Table 1.

**Table 1:** Participant Demographics.

|  | Vignette |  |
| --- | --- | --- |
|  | Bleed (n=652) | Cancer (n=682) |
| Age – Mdn (min-max) | 44 (18-78) | 45 (18-85) |
| Sex – n (%) |  |  |
| Men | 302 (46.9%) | 306 (45.2%) |
| Women | 343 (53.2%) | 371 (54.8%) |
| Race – n (%) |  |  |
| White/Caucasian | 443 (68.6%) | 492 (72.7%) |
| Black/African American | 75 (11.6%) | 72 (10.6%) |
| Asian | 49 (7.6%) | 47 (6.9%) |
| Native Hawaiian/Other Pacific Islander | 2 (0.3%) | 1 (0.2%) |
| Indigenous/Native American | 6 (0.9%) | 9 (1.3%) |
| More than one race | 71 (11.0%) | 56 (8.3%) |
| Household size – n (%) |  |  |
| 1 person | 130 (20.1%) | 148 (22.0%) |
| 2-4 people | 448 (69.4%) | 463 (68.7%) |
| 5+ people | 68 (10.5%) | 63 (9.4%) |
| Household income – Mdn (min-max) | \$68k (\$0-\$600k) | \$68k (\$0-\$875k) |

Note. Mdn=median

### Procedure

Participants provided consent and were directed to an online survey. The survey was comprised of two vignettes and demographic questions. Participants received $2.50 for participation. Both vignettes presented a hypothetical case where a radiologist failed to identify an abnormality and was being sued by the patient or the patient’s family member. Participants, like real jurors, indicated whether the radiologist had met their duty of care to the patient. Generally, in medical malpractice cases where a radiologist has been sued for perceptual error, jurors determine whether the radiologist failed to exercise the degree of care, skill, and judgment that a reasonable doctor would exercise in the same or similar circumstances under the prevailing standards of practice for doctors of that specialty.^6^ In the “bleed” vignette, a radiologist failed to detect bleeding in the brain on a CT scan of a stroke patient, leading to the administration of the blood thinner t-PA. This blood-thinning medication further exacerbated bleeding in the brain and caused the patient to suffer irreversible brain damage. In the “cancer” vignette, a radiologist failed to detect an abnormality on a CT scan of a patient’s chest, leading to a delay in treatment that eventually led to the patient’s death. In both vignettes, the radiologist was being sued by the patient (bleed vignette) or the patient’s family (cancer vignette). Moreover, in both vignettes, imaging was shown to the participants, and conflicting expert witness testimony by both the plaintiff and defense was described. The full vignettes are presented in Appendix A (1: brain bleed, 2: cancer).

### Vignette Manipulations and Details

Each participant was randomly assigned to one of five conditions. The control condition had no mention of AI. In the remaining four conditions, participants were told that an AI system was used to assist the radiologist in checking for the presence of an abnormality. Participants were further told that the AI either found pathology (disagreed with the radiologist) or failed to find pathology (agreed with the radiologist). In two of the five conditions, participants were further provided with detail about the AI system. In the case of AI disagree, this was a false discovery rate (FDR) (false alarm), and in the case of AI agree, this was a false omission rate (FOR) (false miss). Thus, the five conditions were as follows:

1. No AI (control)
2. AI finds pathology (AI disagree)
3. AI fails to find pathology (AI agree)
4. AI finds pathology and AI has a FDR of 50% (AI disagree + FDR)
5. AI fails to find pathology and AI has a FOR of 1% (AI agree + FOR).

### Measures

#### Culpability

Radiologist (defendant) culpability was assessed with a single yes/no question: “Did the radiologist meet their duty of care to the patient?” A response of “no” indicates siding with the plaintiff; a response of “yes” indicates siding with the defendant. Prior to answering this item, participants were told “Radiologists, as doctors, owe a duty of care to the patients for whom they are reviewing imaging. This means that, when they review images, radiologists must use the level of care, skill, and knowledge ordinarily used by radiologists under the circumstances of the case you just read about.”

#### Attention and Manipulation Checks

Two attention-check items were included to verify participants’ understanding of the vignette content (e.g. “What was the cause of the patient’s death?”). Two manipulation-check items were included to verify participants understood the manipulations (e.g. “What was the AI system’s conclusion about the patient’s CT scan?”).

### Statistical analyses

All analyses were conducted using SAS Software (SAS Inc., Cary, NC) with the GLIMMIX procedure. The five experimental conditions were examined using generalized linear modeling assuming a binary distribution. Alpha was established a priori at the 0.05 level and all interval estimates were calculated for 95% confidence.

## Results

### Vignette 1 (Brain Bleed)

#### AI Effect

As seen in Supplemental Table 1 and Figure 1, participants who received the **no AI** condition (control) sided with the plaintiff 56.3% of the time. When **AI Agreed** with the radiologist (also missing brain bleed), participants sided with the plaintiff 50.0% of the time, thus showing no evidence that AI agreeing with the radiologist helped or hurt the radiologist’s defense (50.0% vs. 56.3%, p= 0.309). Conversely, when **AI Disagreed** with the radiologist by flagging a brain bleed, participants sided with the plaintiff more often (72.9% of the time), thus showing the AI result hurt the radiologists’ defense compared to no AI (72.9% vs. 56.3%, p= 0.0054). Finally, participants sided with the plaintiff more often when **AI Disagreed** versus when **AI Agreed** with the radiologist (72.9% vs. 50.0%, respectively, p= 0.0001).

**Figure 1:**
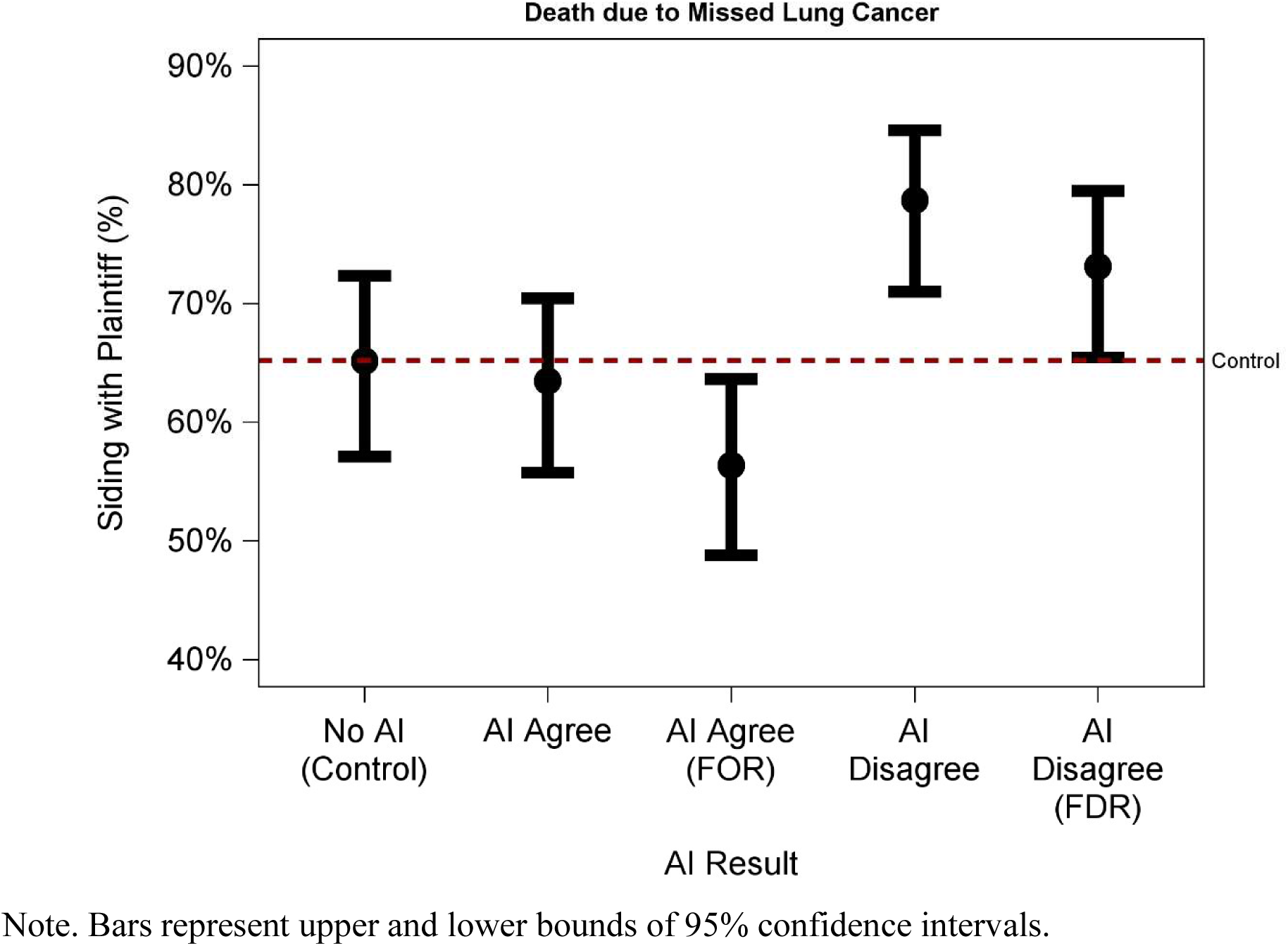
Percent Siding with Plaintiff by Condition – Brain Bleed Note. Bars represent upper and lower bounds of 95% confidence intervals.

#### Context Effect

When **AI Agreed** with the radiologist in also missing the brain bleed, participants sided with the plaintiff 50.0% of the time. However, when participants were provided the false omission rate **(FOR)** of the AI, only 34.0% of participants sided with the plaintiff, thus showing evidence **that including the FOR when AI Agreed with the radiologist helped the radiologist’s defense (**i.e. decreased the percentage who sided with the plaintiff) compared to when **AI agreed** and no FOR was presented (34.0% vs. 50.0%, p=0.0044) and compared to no AI (34.0% vs. 56.3%, p= 0.0003).

Conversely, when **AI Disagreed** with the radiologist by flagging a brain bleed, participants sided with the plaintiff 72.9% of the time. However, when participants were told the false discovery rate **(FDR)** of the AI, only 48.8% of participants sided with the plaintiff, thus showing evidence **that including the FDR when AI Disagreed with the radiologist helped the radiologist’s defense (**i.e. decreased the percentage who sided with the plaintiff) compared to when **AI Disagreed** and no FDR was presented (48.8% vs. 72.9%, p= 0.00005) and even compared to the **no AI control** (48.8% vs. 56.3%, p=0.2288), though this comparison failed to reach statistical significance.

### Vignette 2 (Lung Cancer)

#### AI Effect

As seen in Supplemental Table 1 and Figure 2, participants who received the **no AI** condition (control) sided with the plaintiff 65.2% of the time. When **AI Agreed** with the radiologist in (also missing the lung cancer) participants sided with the plaintiff 63.5% of the time, thus showing no evidence that AI agreeing with the radiologist helped or hurt the radiologist’s defense (63.5% vs. 65.2%, p=0.7678). Conversely, when **AI Disagreed** with the radiologist by flagging a lung cancer, participants sided with the plaintiff more often (78.7% of the time), thus showing the AI result hurt the radiologists’ defense compared to no AI (78.7% vs. 65.2%, p=0.00895). Finally, participants sided with the plaintiff more often when **AI Disagreed** versus when **AI Agreed** with the radiologist (78.7% vs. 63.5%, respectively, p=0.00365).

**Figure 2:**
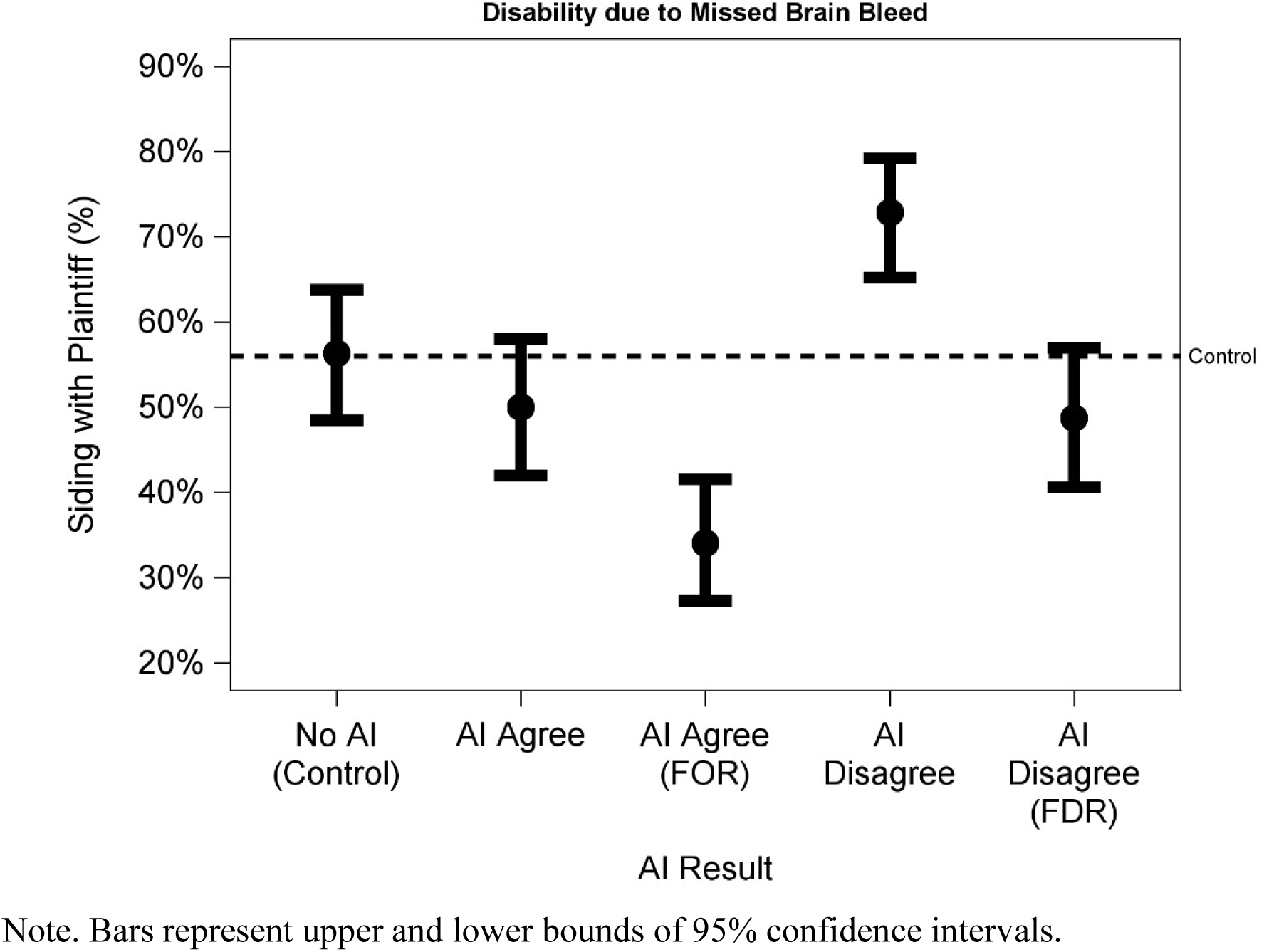
Percent Siding with Plaintiff by Condition – Lung Cancer Note. Bars represent upper and lower bounds of 95% confidence intervals.

#### Context Effect

As noted, when **AI Agreed** with the radiologist in also missing a lung cancer, participants sided with the plaintiff 63.5% of the time. However, when participants were provided the false omission rate **(FOR)** of the AI, only 56.4% of participants sided with the plaintiff, thus showing evidence that **that including the FOR when AI Agreed with the radiologist helped the radiologist’s defense (**i.e. decreased the percentage who sided with the plaintiff) compared to when **AI agreed** and no FOR was presented (63.5% vs. 56.4%, p=0.1085) and compared to **no AI** (56.4% vs. 63.5%, p=0.1342), though both failed to reach statistical significance.

Conversely, when **AI Disagreed** with the radiologist by flagging a lung cancer, participants sided with the plaintiff 78.7% of the time. However, when participants were provided the false discovery rate **(FDR)** of the AI, only 73.1% of participants sided with the plaintiff, thus showing evidence **that including the FDR when AI Disagreed with the radiologist helped the radiologist’s defense (**i.e. decreased the percentage who sided with the plaintiff) compared to when **AI Disagreed** and no FDR was presented (78.7% vs. 73.1%, p=0.1507) and even compared to the **no AI control** (73.1% vs. 65.2%, p=0.16), though both comparisons failed to reach statistical significance.

## Discussion

Legal scholars are beginning to consider how artificial intelligence might change the nature of medical malpractice liability in radiology.^7^ Mello and Guha^8^ discuss difficulties in applying existing tort law to such cases and indicate that it may take time for the relevant legal precedents to become more fully established. Nonetheless, we can be confident that there will eventually be cases in which a radiologist who has used an AI system is being sued for malpractice and jurors consider whether that radiologist has agreed or disagreed with the system as they determine whether the standard of care was met.^9^

We found that participants were more likely to side with the plaintiff, finding that a radiologist had failed to meet their duty of care, when told the radiologist *disagreed* with an AI system. Put differently, when AI found a pathology but the radiologist failed to find any abnormality, radiologists were judged as *more culpable* compared to instances where AI also indicated a case was normal (AI agree condition), or no AI was used (no AI condition). The latter comparison – AI disagree vs. no AI – is especially relevant as it shows the increased liability of missing a pathology when AI finds a pathology relative to standard of care (no AI).

Our study suggests that radiologists will be judged more harshly in a courtroom setting when disagreeing with an AI system that determines a pathology is present. It is important to understand the likely consequences of this. Due to “defensive medicine biases”^5^ radiologists will be incentivized to call cases as abnormal that AI calls as abnormal even when they strongly believe AI is incorrect. Left unresolved, this has the potential to exacerbate our current problem of over-diagnosis and over-testing.^10^

Moreover, our study suggests that using AI is a one-way ratchet in favor of finding liability: disagreeing with AI appears to increase liability risk, but agreeing with AI fails to decrease liability risk relative to not using AI. In this respect, it is fair to characterize the effect of AI use as a potential “AI penalty” in litigation for perceptual errors.

Fortunately, however, our study suggests there is an important strategy for minimizing the AI penalty. When participants were told that 50% of cases that AI indicated were positive were actually negative (i.e. 50% FDR), they were more likely to side with the radiologist compared to when no FDR was provided. The magnitude of this effect was stark in the brain bleed vignette (i.e. 48.8% vs. 72.9% of participants sided with the plaintiff in the AI disagree and AI disagree + FDR conditions, respectively) and mild in the lung cancer vignette (i.e. 73.1% vs. 78.7%) where it failed to reach statistical significance even though the directionality of the effect was consistent.

The cause of the difference between vignettes cannot be established with the current study, though two possibilities are the different pathologies (cancer vs. brain bleed) and different patient outcomes (death vs. disability). Importantly, however, for both vignettes, the difference between no AI and AI disagree + FDR was not statistically significant. This means we did not find evidence that radiologists were viewed as more liable when they disagreed with AI and the FDR was known compared to standard of care.

Finally, when AI agreed with the radiologist, providing a FOR also decreased judgements against the radiologist, though this was also only significant for the brain bleed vignette. Our findings suggest that whether a radiologist agrees or disagrees with AI, a radiologist’s liability is reduced when the AI’s error rate is discussed. The fact we observed similar findings for AI disagree + FDR vs. AI disagree and AI agree + FOR vs. AI agree suggests that priming people with the notion of (stochastic) imperfection in one entity (AI) may make jurors less likely to find that radiologists breached the standard of care for perceptual errors.

## Limitations

The information shown to participants was much less detailed than what would be seen by a real jury, which limits ecological validity. However, effects observed in the context of a brief vignette study where people are less motivated to carefully process information than they would be if on a jury, suggests that in the real-world the magnitude of these effects might be even larger. Another challenge to ecological validity is that in a courtroom setting jurors deliberate together, whereas in our study participants responded to questions by themselves. Again, we believe that this also likely dampens the magnitude of our effects, as information used for the experimental manipulations (e.g., presence of AI data) could be made salient during deliberation by members of the jury who viewed those facts as relevant for other jury members who did not initially view such information as important. Our study sampled a broad range of adult U.S. citizens to approximate the population of potential jurors. However, Prolific participants are likely not representative of American citizens as a whole. Moreover, the process of voir dire excuses many people from serving on real cases, which cannot be mimicked using our survey methodology. In addition, our results are conditional: AI (theoretically) should prevent cases from being missed, so overall, liability should be diminished. However, in the event when a case is missed and AI was used, then relative liability may increase in the case where AI detected something relative to no AI or AI also missed it. Finally, both of our cases were similar in that a radiologist was being sued for missing an abnormality and results may not translate to other types of cases. However, failure to diagnose is by far the most common type of lawsuit that radiologists face.^11^

To our knowledge, this study is the first empirical examination of how people react to differing AI information when presented with a hypothetical case of a radiologist being sued. Our results suggest that radiologists may be penalized in a courtroom if they miss an abnormality that was detected by an AI system. However, this can be mitigated or corrected by presenting data regarding AI precision. These findings are relevant for radiologists, patients, and attorneys.

## Data Availability

All data produced in the present study are available upon reasonable request to the authors

## Appendix A Vignettes

### 1. Brain Bleed Vignettes

A 50-year-old with acute neurological symptoms visited the Emergency Department of a hospital. The neurological symptoms were signs of a possible stroke.

The doctor wanted to treat the patient with a powerful blood thinner called t-PA. However, prior to administration of t-PA, a computerized tomography (CT) scan of the brain (Scan 1) was obtained. A CT is a type of x-ray imaging. The CT was performed to ensure there was no bleeding of the brain present, **as giving a patient t-PA would worsen brain bleeding.**

**No AI (Control):**

Radiologist Y (radiologists are doctors who specialize in reading images) interpreted the CT as **no evidence of bleeding in the brain** and therefore, the blood thinner, t-PA, was administered.

**AI Disagree:**

An Artificial Intelligence (AI) system was used by Radiologist Y’s practice (radiologists are doctors who specialize in reading images). The AI system is designed to examine CT scans for brain bleeding. The AI system flagged the patient’s case, which means **AI identified it as a brain bleed**. Knowing that AI flagged the patient’s case, Radiologist Y then also interpreted the CT. However, Radiologist Y **disagreed** with the AI system and instead concluded that there was **no evidence of bleeding in the brain** and therefore, the blood thinner, t-PA, was administered.

**AI Agree:**

An Artificial Intelligence (AI) system was used by Radiologist Y’s practice (radiologists are doctors who specialize in reading images). The AI system is designed to examine CT scans for brain bleeding. The AI system did not flag the patient’s case, which means **AI found no evidence of a brain bleed**. Knowing that AI did not flag the patient’s case, Radiologist Y then also interpreted the CT. Radiologist Y **agreed** with the AI system and also concluded that there was **no evidence of bleeding in the brain** and therefore, the blood thinner, t-PA, was administered.

**AI Disagree + FDR:**

An Artificial Intelligence (AI) system was used by Radiologist Y’s practice (radiologists are doctors who specialize in reading images). The AI system is designed to examine CT scans for brain bleeding. However, no AI system is perfect. This particular AI system has a “false alarm” rate of 50%. Thus, for every 10 cases that AI flags as a brain bleed, only 5 are actual brain bleeds. The AI system flagged the patient’s case, which means **AI identified it as a brain bleed**. Knowing that AI flagged the patient’s case, Radiologist Y then also interpreted the CT. However, Radiologist Y **disagreed** with the AI system and instead concluded that there was **no evidence of bleeding in the brain** and therefore, the blood thinner, t-PA, was administered.

**AI Agree + FOR:**

An Artificial Intelligence (AI) system was used by Radiologist Y’s practice (radiologists are doctors who specialize in reading images). The AI system is designed to examine CT scans for brain bleeding. However, no AI system is perfect. This particular AI system has a “miss” rate of 1%. Thus, for every 100 cases that AI finds no evidence of a brain bleed, it misses 1 case that actually is a brain bleed. The AI system did not flag the patient’s case, which means **AI found no evidence of a brain bleed**. Knowing that AI did not flag the patient’s case, Radiologist Y then also interpreted the CT. Radiologist Y **agreed** with the AI system and also concluded that there was **no evidence of bleeding in the brain** and therefore, the blood thinner, t-PA, was administered.

After getting t-PA, the patient’s symptoms got much worse, so another CT scan was taken (Scan 2). This new scan (Scan 2) showed a very large brain bleed.

**Scan 1:**
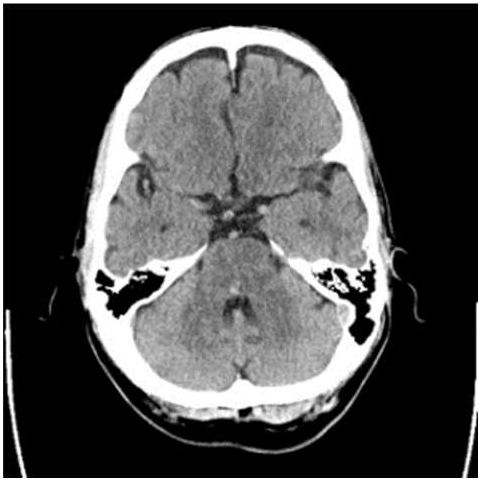
The original CT scan.

**Scan 2:**
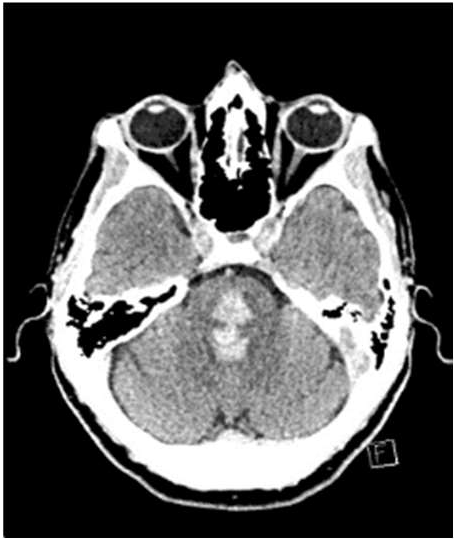
Repeat CT scan.

The plaintiff for this case is the patient and the defendant is Radiologist Y. The missed brain bleed on the original CT scan (Scan 1) was the direct cause of the stroke becoming worse because: 1) t-PA caused extra bleeding and 2) t-PA was only given because Radiologist Y did not see the brain bleed on Scan 1. As a result of the miss, the patient suffered irreversible brain damage and the magnitude of their permanent disability is far greater than what would have occurred had the brain bleed been successfully detected on Scan 1 by Radiologist Y.

At trial, the plaintiff called an independent radiologist as an expert witness who stated the brain bleed on the CT scan was obvious and should have been detected. This person received **$11,000** as compensation for serving as an expert witness.

The defense called their own independent radiologist as an expert witness. **Upon reading the image, the defense expert witness, like Radiologist Y, did not detect the brain bleed.** When later shown the brain bleed, the defense expert witness concluded that it was very subtle and that most radiologists would not have seen it. This person received **$4,500** as compensation for serving as an expert witness.

Note. Vignettes were identical across conditions aside from the differences outlined above.

### 2. Cancer Vignettes

A 55-year-old woman with a history of breast cancer 9 years earlier underwent a routine screening computerized tomography (CT) scan of the chest. A CT scan is a type of x-ray imaging.

**No AI (Control):**

Radiologist A (radiologists are doctors who specialize in reading images) interpreted the CT as **no evidence of abnormality** even though there was actually a soft tissue mass on the left anterior chest wall, which could indicate cancer.

**AI Disagree:**

An Artificial Intelligence (AI) system was used by Radiologist A’s practice (radiologists are doctors who specialize in reading images). The AI system is designed to examine CT scans for abnormalities. The AI system flagged the patient’s case, which means **AI identified it as abnormal**. Knowing that AI flagged the patient’s case, Radiologist A then also interpreted the CT. However, Radiologist A **disagreed** with the AI system and instead concluded that there was **no evidence of abnormality** even though there was actually a soft tissue mass on the left anterior chest wall, which could indicate cancer.

**AI Agree:**

An Artificial Intelligence (AI) system was used by Radiologist A’s practice (radiologists are doctors who specialize in reading images). The AI system is designed to examine CT scans for abnormalities. The AI system did not flag the patient’s case, which means **AI found no evidence of abnormality**. Knowing that AI did not flag the patient’s case, Radiologist A then also interpreted the CT. Radiologist A **agreed** with the AI system and also concluded that there was **no evidence of abnormality** even though there was actually a soft tissue mass on the left anterior chest wall, which could indicate cancer.

**AI Disagree + FDR:**

An Artificial Intelligence (AI) system was used by Radiologist A’s practice (radiologists are doctors who specialize in reading images). The AI system is designed to examine CT scans for abnormalities. However, no AI system is perfect. This particular AI system has a “false alarm” rate of 50%. Thus, for every 10 cases that AI flags as abnormal, only 5 are actually abnormal. The AI system flagged the patient’s case, which means **AI identified it as abnormal**. Knowing that AI flagged the patient’s case, Radiologist A then also interpreted the CT. However, Radiologist A **disagreed** with the AI system and instead concluded that there was **no evidence of abnormality** even though there was actually a soft tissue mass on the left anterior chest wall, which could indicate cancer.

**AI Agree + FOR:**

An Artificial Intelligence (AI) system was used by Radiologist A’s practice (radiologists are doctors who specialize in reading images). The AI system is designed to examine CT scans for abnormalities. However, no AI system is perfect. This particular AI system has a “miss” rate of 1%. Thus, for every 100 cases that AI finds no evidence of a brain bleed, it misses 1 case that actually is a brain bleed. The AI system did not flag the patient’s case, which means **AI found no evidence of abnormality**. Knowing that AI did not flag the patient’s case, Radiologist A then also interpreted the CT. Radiologist A **agreed** with the AI system and also concluded that there was **no evidence of abnormality** even though there was actually a soft tissue mass on the left anterior chest wall, which could indicate cancer.

One year later, the patient began to have pain in her chest, and a biopsy was ultimately performed. A biopsy is when a tissue sample is taken and analyzed. The biopsy revealed metastatic disease, which means it was actually cancer. The cancer had spread to other parts of her body.

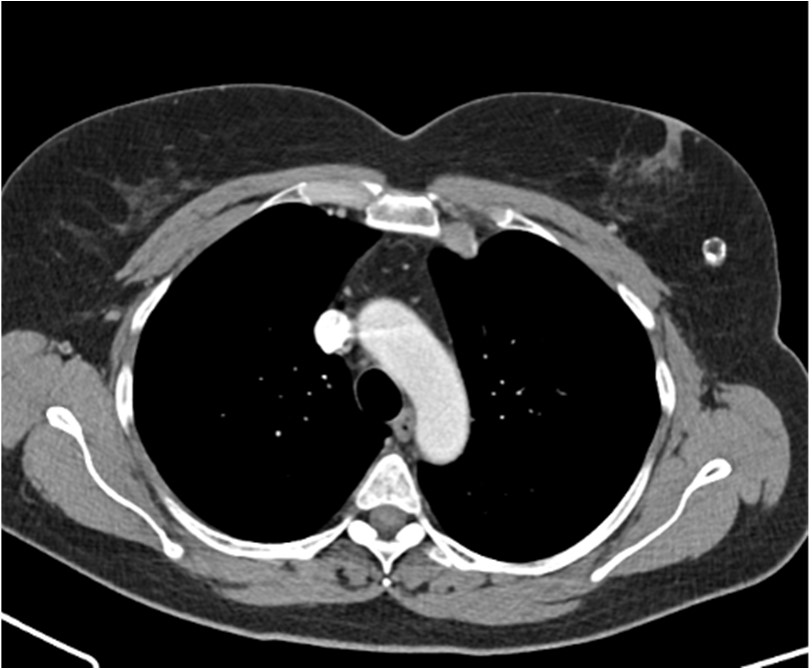
The image below is from the original CT scan.

One year later, the patient died due to cancer. The plaintiff for this case is the patient’s family and the defendant is Radiologist A. The one-year delay in the diagnosis of cancer caused by Radiologist A missing the finding on the CT screen significantly reduced her chances of survival.

At trial, the plaintiff called an independent radiologist as an expert witness who stated the abnormality on the CT scan was obvious and should have been detected. This witness received **$12,000** as compensation for serving as an expert witness.

The defense called their own independent radiologist as an expert witness. **Upon reading the image, the defense expert witness, like the radiologist being sued (Radiologist A), also did not detect the abnormality.** When later shown the abnormality, the defense expert witness concluded that it was very subtle and most radiologists would not see it. This witness received **$5,000** as compensation for serving as an expert witness.

Note. Vignettes were identical across conditions aside from the differences outlined above.

---

**Supplemental Table 1:**
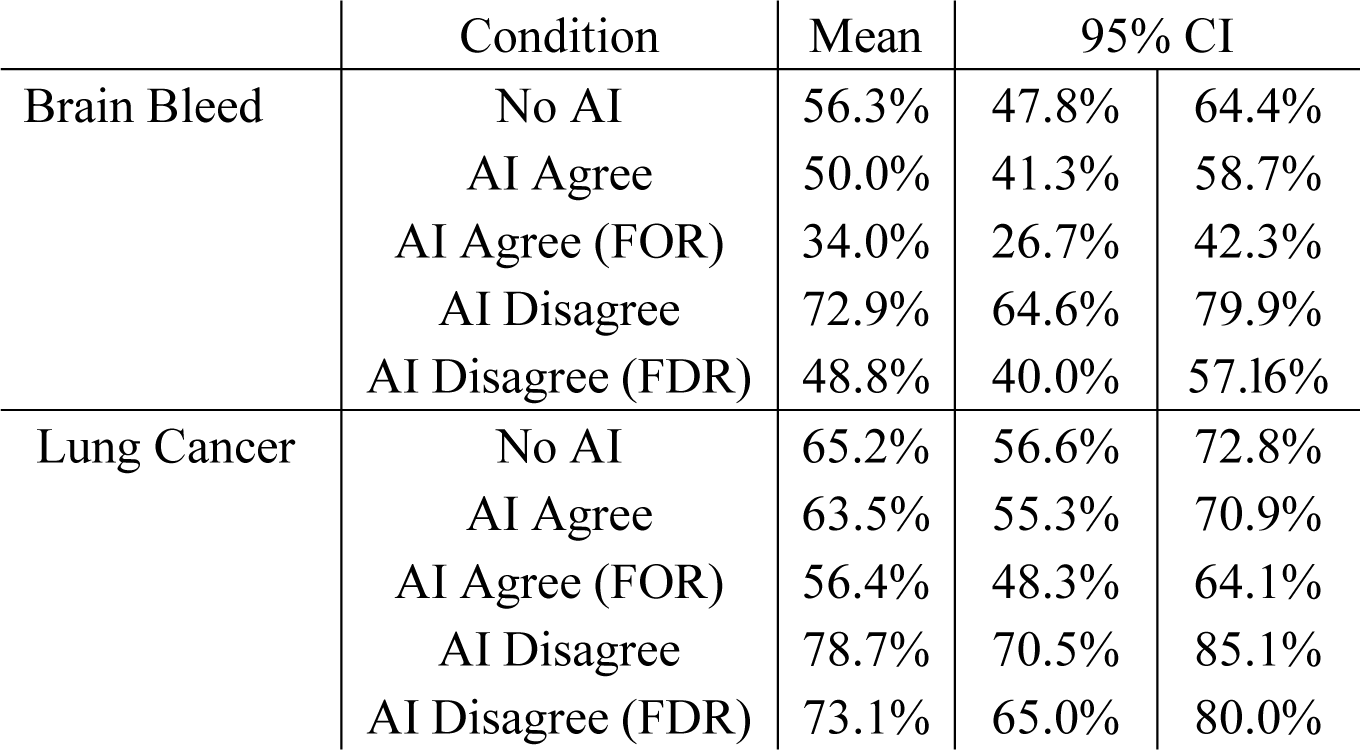
Percent Siding with Plaintiff by Condition – Brain Bleed and Lung Cancer Vignettes.

|  | Condition | Mean | 95% CI |  |
| --- | --- | --- | --- | --- |
| Brain Bleed | No AI | 56.3% | 47.8% | 64.4% |
|  | AI Agree | 50.0% | 41.3% | 58.7% |
|  | AI Agree (FOR) | 34.0% | 26.7% | 42.3% |
|  | AI Disagree | 72.9% | 64.6% | 79.9% |
|  | AI Disagree (FDR) | 48.8% | 40.0% | 57.16% |
| Lung Cancer | No AI | 65.2% | 56.6% | 72.8% |
|  | AI Agree | 63.5% | 55.3% | 70.9% |
|  | AI Agree (FOR) | 56.4% | 48.3% | 64.1% |
|  | AI Disagree | 78.7% | 70.5% | 85.1% |
|  | AI Disagree (FDR) | 73.1% | 65.0% | 80.0% |

